# The effect of D-cycloserine on brain processing of breathlessness over pulmonary rehabilitation - an experimental medicine study

**DOI:** 10.1101/2021.06.24.21259306

**Authors:** Sarah L. Finnegan, Olivia K. Harrison, Sara Booth, Andrea Dennis, Martyn Ezra, Catherine J. Harmer, Mari Herigstad, Bryan Guillaume, Thomas E. Nichols, Najib M. Rahman, Andrea Reinecke, Olivier Renaud, Kyle T.S. Pattinson

## Abstract

**Background:** Pulmonary rehabilitation is the best treatment for chronic breathlessness in COPD but there remains an unmet need to improve efficacy. Pulmonary rehabilitation has strong parallels with exposure-based cognitive behavioural therapies (CBT), both clinically and in terms of brain activity patterns. The partial NMDA-receptor agonist, D-cycloserine has shown promising results in enhancing efficacy of CBT, thus we hypothesised that it would similarly augment the effects of pulmonary rehabilitation in the brain. Positive findings would support further development in phase 3 clinical trials.

**Methods:** 72 participants with mild-to-moderate COPD were recruited to a double-blind pre-registered (ID: NCT01985750) experimental medicine study running parallel to a pulmonary rehabilitation course. Participants were randomised to 250mg D-cycloserine or placebo, administered immediately prior to the first four sessions of pulmonary rehabilitation. Primary outcome measures were differences between D-cycloserine and placebo in brain activity in the anterior insula, posterior insula, anterior cingulate cortices, amygdala and hippocampus following completion of pulmonary rehabilitation. Secondary outcomes included the same measures at an intermediate time point and voxel-wise difference across wider brain regions.

**Results:** No difference between D-cycloserine and placebo groups was observed across the primary or secondary outcome measures. Questionnaire and measures of respiratory function showed no group difference.

**Conclusions:** This is the first study testing brain-active drugs in pulmonary rehabilitation. Rigorous trial methodology and validated surrogate end-points maximised statistical power. Although increasing evidence supports therapeutic modulation of NMDA pathways to treat symptoms, we conclude that a phase 3 clinical trial of D-cycloserine would not be worthwhile.

**Key Messages:** *What is the key question?:* Does the partial NMDA-receptor agonist, D-cycloserine, augment the effects of pulmonary rehabilitation on breathlessness related brain activity?

*What is the bottom line?:* Rigorous trial methodology and validated surrogate end-points revealed no effect of D-cycloserine on breathlessness related brain activity across pulmonary rehabilitation.

*Why read on?:* This study highlights both the value of functional magnetic resonance imaging in “de-risking” expensive clinical trials and provides detailed investigation of brain-targeted points for pharmacological treatments of breathlessness.

## Introduction

Chronic breathlessness is a central symptom of chronic obstructive pulmonary disease (COPD). Currently, pulmonary rehabilitation offers the most effective treatment strategy for chronic breathlessness in COPD. However, around 30% of patients derive no clinical benefit [1]. Health-related benefits plateau within the first 6-months following pulmonary rehabilitation, returning to pre-rehabilitation levels for the majority of patients after 12-18 months [2]. Thus, there remains an unmet need to develop strategies to increase or prolong the beneficial effects of pulmonary rehabilitation.

A body of evidence has shown that improvements in breathlessness over pulmonary rehabilitation result from a reappraisal of the sensory experience [3, 4], arresting the downward spiral of fear, avoidance and physical deconditioning. The safe, graded exposure to breathlessness within pulmonary rehabilitation parallels techniques of exposure-based cognitive behavioural therapy (CBT), in which pathological fears are first activated and then disconfirmed by new adaptive information. These similarities are highted by clinical studies of both pulmonary rehabilitation and cognitive behavioural therapy, which show changes to brain activity within areas associated with attention and learned sensory and emotional expectations such as cingulate cortex, angular gyrus, insula and supramarginal gyrus, [4, 5].

In the field of psychiatry, there has been great interest in the partial NMDA agonist D-cycloserine, as a pharmacological adjunct to enhance efficacy of exposure-based CBT [6-8]. D-cycloserine is thought to act at the glycine modulatory site of the NMDA receptor. Its high affinity binding enhances synaptic plasticity, promoting emotional learning processes [8] boosting therapeutic effects of CBT as a result [7, 9, 10]. Experimental medicine studies have demonstrated reductions in emotional response within the amygdala when paired with CBT [6], increasing activity within hippocampus in a manner linked to learning [11]. In CBT for alcoholism, D-cycloserine decreased cue induced brain activity across the ventral and dorsal striatum which was associated with reductions in alcohol craving [12]. Clinical trials of D-cycloserine paired with CBT have demonstrated reductions in symptoms of acrophobia [13], social phobia [14], panic disorder [15] and obsessive compulsive disorder (OCD) over placebo [16, 17], often with medium to large effect sizes.

Given the strong parallels between pulmonary rehabilitation and exposure-based CBT, and the strong effects of pulmonary rehabilitation on affective components of breathlessness [3, 4] we hypothesised that D-cycloserine may have therapeutic benefits in enhancing pulmonary rehabilitation.

To test this hypothesis, we performed an experimental medicine study using functional magnetic resonance imaging (FMRI) markers of drug efficacy in the brain. We chose this approach over a phase 3 clinical trial because the differences between COPD populations and those with primary psychiatric conditions in which D-cycloserine has been tested so far might necessitate a bespoke trial design. Therefore, we wanted to test efficacy in the brain first. FMRI end-points are more sensitive than clinical ones [18] and thus facilitate the adoption of more statistically robust approaches. This means that results can be obtained with smaller sample sizes than in a phase 3 clinical trial. A positive result from an FMRI study would facilitate clinical trial design and help early decisions of go/no-go on further clinical development.

## Methods and Materials

An overview of the methodology is presented here. Full details, including non-completion and sensitivity analysis can be found within supplementary materials. The study and statistical analysis plan were pre-registered on clinicaltrials.gov (ID: NCT01985750) prior to unblinding. This is the first example of pre-registration of both study design and analysis plan in a respiratory neuroimaging study.

### Sample Size

At the time of study inception (and to a large extent still to date), the literature regarding D-cycloserine’s effects on functional brain activity is very limited. Therefore, in order to calculate the sample sizes required for this study we first took into account the described effects of D-cycloserine in clinical studies of augmentation for cognitive behavioural therapy for anxiety disorders, where effect sizes of up to 1.06 have been reported (although more commonly 0.4 to 0.7) [13, 14, 19]. The most relevant paper (on treatment of snake phobia [10]) demonstrated that effects observed with neuroimaging were more sensitive than behavioural effects, therefore powering for a behavioural outcome measure (breathlessness-anxiety) provided a safe margin and was likely to be sufficiently conservative to detect our measures of interest. This was particularly the case as compared to the relatively blunt nature of behavioural data collection, functional neuroimaging carries considerably more specificity and statistical power. The study was not therefore specifically powered to investigate the clinical effects of D-cycloserine. In our previous study we observed an 11% (SD15%) improvement in breathlessness-related anxiety, measured with our FMRI word task (pre-treatment mean score 38%, post treatment mean score 27%, difference 11%, SD of difference 15%) [4]. Making a conservative assumption, we estimated that D-cycloserine augments this response with an effect size of 0.4. Assuming a similar coefficient of variation we anticipated an 18% (SD24%) improvement in breathlessness-anxiety (i.e. pre-treatment mean score 38%, post treatment mean score 20%, difference 18%, SD of difference 24%). Assuming α=0.05 and power 0.80, then we estimated a sample size of 36 in each group randomised 1:1. As this is a behavioural outcome, we expected this to have sufficient power to detect change in BOLD signalling.

### Participants

91 participants (30 female, median age 70 years; range 46-85 years) with COPD were recruited immediately prior to their enrolment in a National Health Service-prescribed course of pulmonary rehabilitation (full demographic information including non-continuation is shown in Supplementary material Table 1). From this population, 72 participants completed all study visits (18 female, median age 71 years (46-85 years)) (Table 1 and Figure 2). Written informed consent was obtained from all participants prior to the start of the study. Study approval was granted by South Central Oxford REC B (Ref: 118784, Ethics number: 12/SC/0713). Study inclusion criteria were: a diagnosis of COPD and admittance to pulmonary rehabilitation. Exclusion criteria were: inadequate understanding of verbal and written English, significant cardiac, psychiatric (including depression under tertiary care) or metabolic disease (including insulin-controlled diabetes), stroke, contraindications to either D-cycloserine (including alcoholism) or magnetic resonance imaging (MRI), epilepsy, claustrophobia, regular therapy with opioid analgesics or home oxygen therapy.

**Table 1.** Demographic information from the 72 participants who completed all three study visits. Variance is expression either in terms of standard deviation (SD) or interquartile range (IQR) depending on the normality of the underlying data distribution. **BMI** = Body Mass Index, **MRC** = Medical Research Council. **SpO**_**2**_**%** = Peripheral Oxygen saturation, expressed as a percentage. **FEV** = Forced Expiratory Volume. **FVC** = Forced Vital Capacity. Also listed with prevalence in brackets are recorded comorbidities ordered by frequency

| Visit 1<br>(Pre-rehabilitation) |  |  |
| --- | --- | --- |
|  | D-cycloserine | Placebo |
| Age (median years / range) | 71.0 / (47-81) | 71.5 / (46-85) |
| Smoking pack-years (years / IQR) | 34.0 / (25.6) | 30.0 / (30.0) |
| Duration of breathlessness (years / IQR) | 8.0 (17.0) | 9.5 (10.0) |
| Total exacerbations (number / IQR) | 0.0 (1.0) | 1.0 (2.3) |
| BMI (kg.m-2 ± SD) | 27.3 ± 6.5 | 26.9 ± 5.7 |
| MRC breathlessness scale (IQR) | 3.0 (1) | 2.5 (1) |
| Resting SpO<sub>2</sub>% (IQR) | 95 / (3.3) | 95 / (3.0) |
| Resting heart rate (beats.min-1 ± SD) | 80.8 ± 13.4 | 80.8 ± 14.9 |
| FEV1/FVC (IQR) | 0.53 / (0.17) | 0.56 / (0.13) |
| Comorbidities (frequency) |  |  |
|  | Asthma (14) | Asthma (11) |
|  | Hypertension (13) | Hypertension (11) |
|  | Gastro-oesophageal reflux (10) | Gastro-oesophageal reflux (12) |
|  | Swelling of both ankles (11) | Swelling of both ankles (8) |
|  | Surgery to the chest (6) | Surgery to the chest (7) |
|  | Depression (6) | Depression (2) |
|  | Diabetes (3) | Diabetes (6) |
|  | Heart attack (4) | Heart attack (5) |
|  | Bronchiectasis (3) | Bronchiectasis (4) |
|  | Osteoporosis (2) | Osteoporosis (4) |
|  | Arrhythmia (3) | Arrhythmia (4) |
|  | Inflammatory bowel disease (2) | Inflammatory bowel disease (3) |
|  | Peptic ulcer (3) | Peptic ulcer (2) |
|  | Heart failure (1) | Heart failure (1) |
|  | Tuberculosis (1) | Neuromuscular weakness (2) |

### Study drug

Participants were randomised in a double-blinded procedure to receive either 250mg oral D-cycloserine or a matched placebo, administered by the study nurse 30 minutes prior to the onset of their first four pulmonary rehabilitation sessions. 250mg dosage has been shown to be efficacious [6, 11]. D-cycloserine exerts its best effects when given immediately before exposure based therapy and only for a limited number of times [20] with daily administration associated with tachyphylaxis [13]. Therefore, the dose timing of D-cycloserine was deliberately selected to be the first four sessions, where the greatest potential for emotional learning occurs as patients become habituated to pulmonary rehabilitation. Study participants, investigators and those performing the analysis were blinded to the treatment allocation. Both D-cycloserine and placebo were over-encapsulated to appear identical.

The following minimization criteria were used for randomisation a) centre at which pulmonary rehabilitation was performed, MRC breathlessness grade, presence of diabetes, whether the participant was taking antidepressants, the age at which the participant completed full time education and previous pulmonary rehabilitation. Full details of randomisation can be found within the supplementary materials.

### Study visit protocol

Following telephone screening, participants were invited to attend their first research session (baseline) prior to starting pulmonary rehabilitation. A second study visit took place following the fourth pulmonary rehabilitation session but before the sixth session. Participants completed the remainder of their pulmonary rehabilitation course before attending a third study session (Figure 2) that occurred as close to the termination of pulmonary rehabilitation as possible and always within two weeks.

### Pulmonary Rehabilitation

Pulmonary rehabilitation courses were run by either Oxford Health NHS Foundation Trust, West Berkshire NHS Foundation Trust, or Milton Keynes University Hospitals NHS Trust.

### Self-report questionnaires

Building on our previous work [4, 21] we selected a set of questionnaires with proven sensitivity to changes across pulmonary rehabilitation [4], which were designed to probe the experience of living with COPD. These were scored according to their respective manuals: Dyspnoea-12 (D12) Questionnaire [22], Centre for Epidemiologic Studies Depression Scale (CES-D) [23], Trait Anxiety Inventory (TRAIT) [24], Fatigue Severity Scale [25], St George’s Respiratory Questionnaire (SGRQ) [26], Medical Research Council (MRC) breathlessness scale [27], Mobility Inventory (MI) [28], Breathlessness catastrophising scale, adapted from the Catastrophic Thinking Scale in Asthma [29], Breathlessness vigilance, adapted from the Pain Awareness and Vigilance Scale [30].

### Physiological measures

Spirometry and two Modified Shuttle Walk Tests (MSWT) were collected using standard protocols [31, 32]. Participant height and weight were recorded at each visit. Oxygen saturations and heart rate were measured with pulse oximetry and were collected at rest and following the MSWT.

### MRI measures

#### Image acquisition

Magnetic resonance imaging of the brain was carried out using a Siemens 3T MAGNETOM Trio. A T1-weighted (MPRAGE) structural scan (voxel size: 1 × 1 × 1 mm) was collected and used for registration purposes. A T2*-weighted, gradient echo planar image (EPI) scan sequence (voxel size: 3 × 3 × 3 mm), TR, 3000ms; TE 30ms was used to collect FMRI data.

#### Word cue task

To probe the neural responses of breathlessness-related expectations we examined the activity of brain regions responding to breathlessness-related word cues [4, 21, 33]. This paradigm has previously been shown to be sensitive to improvements in breathlessness over a course of pulmonary rehabilitation. Brain activity was correlated with corresponding visual analogue ratings of breathlessness and breathlessness-anxiety collected during scanning. [4]. During FMRI scanning, participants were presented with a word cue, e.g., “climbing stairs” in white text on a black background for 7 seconds. Participants were then asked, “how breathless would this make you feel” (wB) and “how anxious would this make you feel” (wA). To each question participants responded within a 7 second window using a button box and visual analogue scale (VAS). The response marker always initially appeared at the centre of the scale, with the anchors “Not at all” and “Very much” at either end. Scan duration was 7 minutes and 33 seconds.

#### Control task

A validated task of emotional faces was used as a control to separate generalized anxiety from breathlessness specific anxiety. Fearful or happy faces were presented on a black background was used to examine whether any differences in brain activity patterns between D-cycloserine and placebo groups was specific to breathlessness processing. Each face was shown for 500ms in blocks of 30 seconds. A fixation cross was interspersed for 30 seconds between the blocks of faces. Participants were instructed to respond via a button box to indicate facial gender. Reaction time and accuracy were recorded throughout the task. Scan duration was 5 minutes and 42 sections.

### Outcomes

Our primary outcomes focused on brain activity changes within five key regions of interest, identified in previous studies of breathlessness [4, 34]. The regions of the anterior insular cortex, posterior insular cortex, anterior cingulate cortex, amygdala and hippocampus have all been linked to body and symptom perception as well as emotional salience [35, 36]. Focusing on a small number of regions of interest is more statistically powerful and therefore more likely to detect a hypothesised difference. Secondary hypotheses examined the effect of D-cycloserine across a wider region of interest containing fifteen pre-defined brain areas. The fifteen brain areas encompassed regions associated with sensory and affective processing of breathlessness as well as body and symptom perception. regions.

### Analysis

A summary of analyses is outlined here. Full details, including procedures for dealing with missing data and sensitivity analysis can be found within the supplementary materials. Our analysis was pre-registered and made publicly available prior to unblinding https://mfr.osf.io/render?url=https://osf.io/wqyf4/?action=download%26mode=render.

#### Brain imaging analysis

Image processing was carried out using the Oxford Centre for Functional Magnetic Resonance Imaging of Brain Software Library (FMRIB, Oxford, UK; FSL version 5.0.8; https://ww.fmrib.ox.ac.uk/fsl/), MATLAB R2018b (Mathworks, Natick, MA), R-studio, R version 3.6.1 (2019-07-05). and associated custom scripts. Functional MRI processing was performed using FEAT (FMRI Expert Analysis Tool, within the FSL package).

Data were pre-processed according to standard protocols before being entered into single subject general linear models. These models captured brain activity during the periods in which the breathlessness-related word cues were presented allowing us to examine expectation-related processes.

#### Group level analysis

For each patient, the following metrics were extracted from each of the five regions of interest: anterior insula cortex, posterior insula cortex, anterior cingulate cortex, amygdala and hippocampus, at visits one, two and three:

1. Mean brain activity in response to breathlessness word-cue presentation
2. Mean brain activity for control task of emotional faces

To test for a drug effect across each metric, the values from visit two were entered into independent linear mixed effects models where they were adjusted for age, gender and scores at visit one. To correct for multiple comparisons across regions, permutation testing (with Family Wise Error Rate (FWE) 5%) was carried out. This process was repeated separately for data collected at visit three. Models were programmed using the lme4 function and permuco package within R version 3.6.1 (2019-07-05).

To test for a drug effect across the larger region of interest (panel B of Supplementary Figure 1), the following voxel-wise information was collected from within the region of interest at visits one, two and three:

1. Voxel-wise brain activity in response to breathlessness word-cues presentation
2. Voxel-wise brain activity in response the control task of emotional faces

Each of the values from visit two were entered into independent general linear model (GLM), controlling for age, gender and scores at visit one. Permutation testing was performed with threshold free cluster enhancement (TFCE) (a non-parametric test) [37] using FSL’s Randomise tool [38] at family wise error corrected p<0.05. The process was then repeated separately for data collected at visit three.

## Results

Of the 91 participants recruited (Figure 1), 72 participants completed all three study visits. Reasons for drop-out or exclusion included illness, scanner error and issues with data quality. One further participant was excluded due to an error in task-data collection. 71 participants were therefore assessed for study objectives. Sensitivity analysis was performed and is reported within supplementary materials.

**Figure 1.**
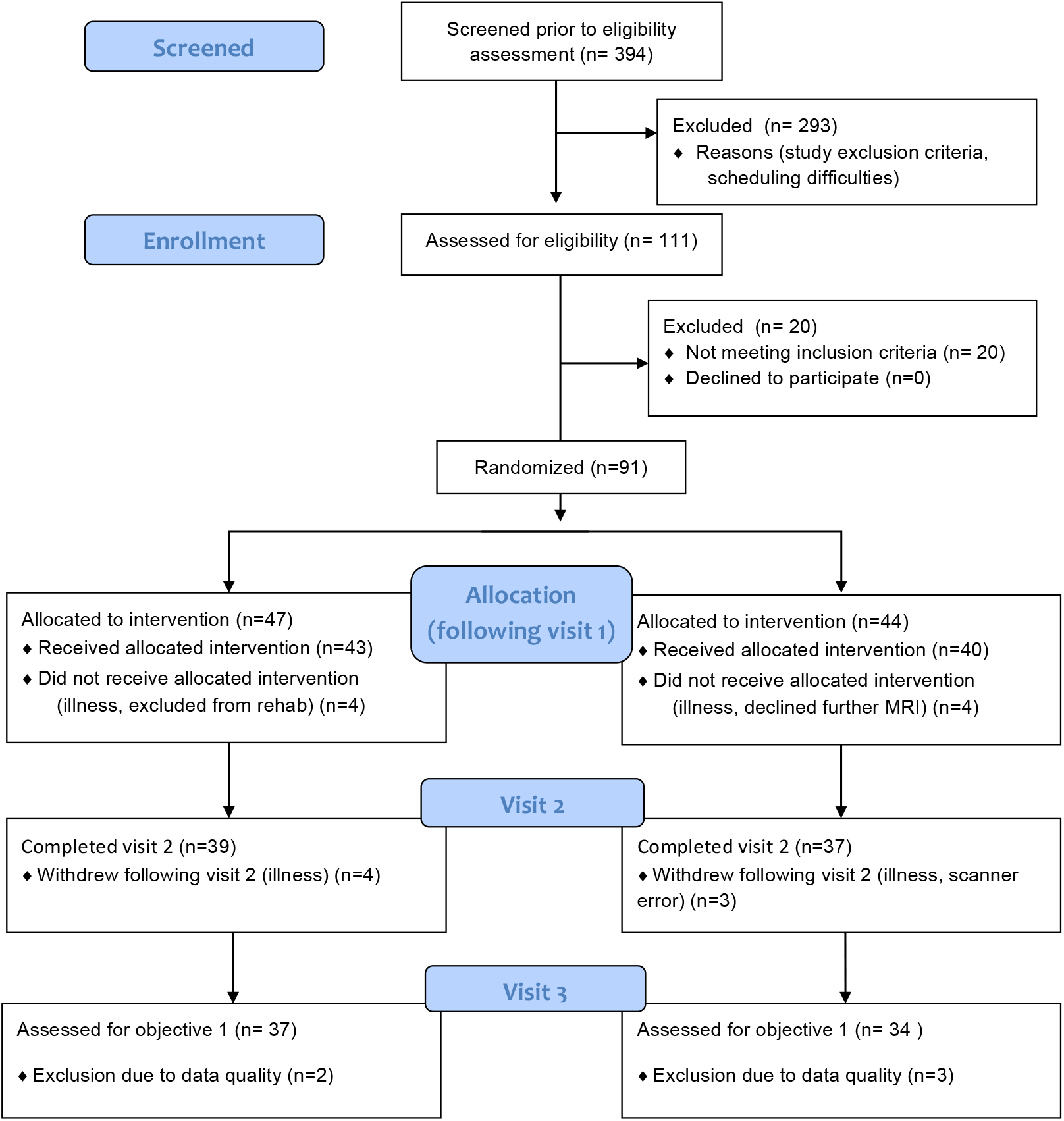
Consort diagram illustrating stages of participant recruitment from initial screening through to completion at visit 3.

**Figure 2.**
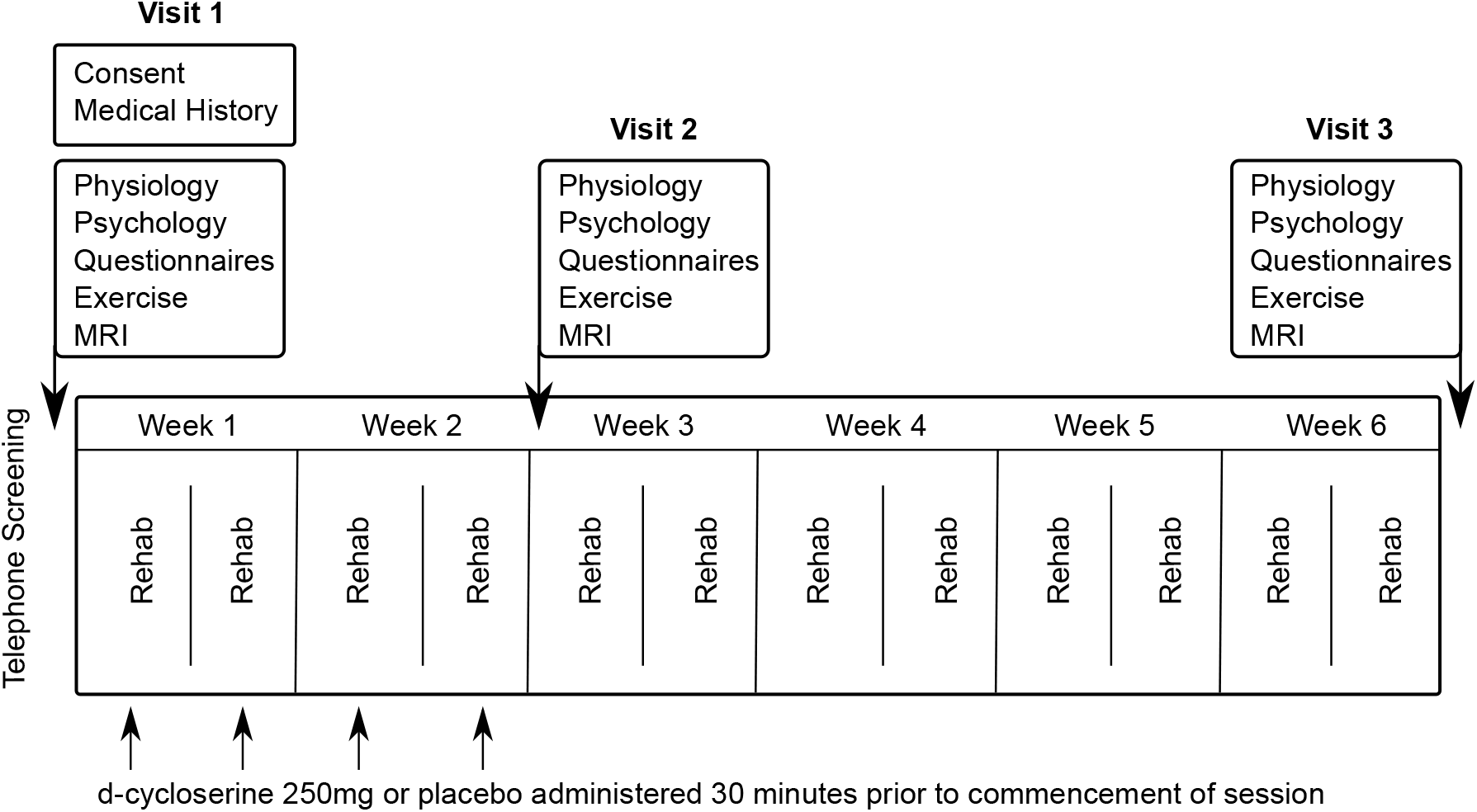
A schematic demonstrating order of visits, rehabilitation sessions and tablet administration throughout the study period. Participants took part in one study visit prior to their first pulmonary rehabilitation session. Study drug/placebo were administered on four occasions over the first four rehabilitation sessions. A second study visit occurred after the final drug/placebo administration. Participants continued with their pulmonary rehabilitation course for a further four weeks before returning for a third study visit.

### Primary outcomes

There was no significant overall effect of D-cycloserine on mean brain activity within the five key regions of interest of anterior cingulate, anterior insula cortex, amygdala, hippocampus or posterior insula cortex (family wise error rate corrected, p>0.05) at visit two or visit three (Table 2).

**Table 2.** Significance of overall effect of D-cycloserine on mean brain activity within the five key regions of interest at visits two, having accounted for scores at visit one. Significance is reported as Family Wise Error (p<0.05) corrected p-values of the difference

| Region of interest | Estimate | Std. Error | Visit two (p-value) |
| --- | --- | --- | --- |
| Anterior cingulate cortex | 0.066 | 0.059 | 0.70 |
| Anterior insula | 0.019 | 0.058 | 0.98 |
| Amygdala | 0.097 | 0.059 | 0.40 |
| Hippocampus | 0.018 | 0.058 | 0.98 |
| Posterior insula | -0.013 | 0.059 | 0.98 |

### Secondary outcomes

There was no significant overall effect of D-cycloserine across the broader mask of fifteen regions measured voxel-wise (family wise error rate correct, p>0.05) at visit two or visit three.

No significant differences in the questionnaire measures or physiology scores were found between the D-cycloserine and placebo groups at any point during the study. No differences were observed between the two groups either in breathlessness ratings (wB) or breathlessness related anxiety (wA) at visit two (both p=0.15 family wise error rate corrected) (Table 3) or visit three (both p=0.053 family wise error rate corrected) (Table 4). No significant effect of drug group was identified, using repeat measured ANOVA’s, for the emotional faces control task (collected during FMRI scanning) at visit two (F(1,68) = 0.17, p=0.68) or visit three (F(1,68) = 0.001, p=0.97). Furthermore, no significant interaction between drug group and emotional valence (happy or fearful faces) was identified at visit two (F(1,68) = 0.36, p=0.55) (Table 4) or visit three (F(1,68) = 0.002, p=0.97) (Table 5). Raw scores are reported in Supplementary Table 4.

**Table 3.** Significance of overall effect of D-cycloserine on mean brain activity within the five key regions of interest at visits three, having accounted for scores at visit one. Significance is reported as Family Wise Error (p<0.05) corrected p-values of the difference

| Region of interest | Estimate | Std. Error | Visit three (p-value) |
| --- | --- | --- | --- |
| Anterior cingulate cortex | -0.009 | 0.056 | 0.98 |
| Anterior insula | 0.019 | 0.055 | 0.98 |
| Amygdala | 0.055 | 0.056 | 0.78 |
| Hippocampus | 0.014 | 0.056 | 0.98 |
| Posterior insula | 0.068 | 0.056 | 0.70 |

**Table 4.** Scores on questionnaire, behavioural and physiological measures for drug and placebo group during (visit two). Measures are expressed as a “change score”. Values are calculated as baseline visit (visit one) minus the later visit, thus positive values indicate an improvement or, in the case of the faces task, a faster reaction time (ms). Significance is reported as Family Wise Error (p<0.05) corrected p-values of the difference in scores between drug and placebo groups at visit two, accounting for scores at visit one. Values are either recorded as mean and standard deviation (1) or as median and interquartile range (2) depending on the normality of distribution. **BMI** = Body Mass Index, **MRC** = Medical Research Council clinical measure of breathlessness, **MSWT =** Modified Shuttle Walk Test, **HR** = Heart rate, **Sats** = Peripheral Oxygen saturation, expressed as a percentage

|  | Visit Two |  |  |
| --- | --- | --- | --- |
|  | D-cycloserine | Placebo | p-value |
| Catastrophising | 1.0 (5.3)<sup>2</sup> | 2.0 (4.0)<sup>2</sup> | 0.84 |
| Depression | 1.2 ± 6.4<sup>1</sup> | 3.2 ± 5.4<sup>1</sup> | 0.55 |
| D12 | 3.8 ± 5.4<sup>1</sup> | 2.2 ± 4.7<sup>1</sup> | 0.55 |
| Fatigue | 7.0 (12.3)<sup>2</sup> | 4.5 (12.0)<sup>2</sup> | 0.64 |
| BMI | -0.16 (0.6)<sup>2</sup> | 0.0 (0.6)<sup>2</sup> | 0.55 |
| MRC | 0 (0.5)<sup>2</sup> | 0 (0.0)<sup>2</sup> | 0.85 |
| MSWT Distance (m) | 0.0 (111.3)<sup>2</sup> | 5.0 (70)<sup>2</sup> | 0.85 |
| MSWT Borg Start | 0 (1)<sup>2</sup> | 0 (1)<sup>2</sup> | 0.85 |
| MSWT Borg Change | 0.5 (1.5)<sup>2</sup> | 0.5 (2)<sup>2</sup> | 0.85 |
| MSWT HR Start | -0.30 ± 13.9<sup>1</sup> | 1.4 ± 13.2<sup>1</sup> | 0.85 |
| MSWT HR Change | -45.4 ± 23.0<sup>1</sup> | -47.5 ± 25.4<sup>1</sup> | 0.90 |
| MSWT Sats Start | -0.30 ± 2.2<sup>1</sup> | -0.1 ± 2.2<sup>1</sup> | 0.85 |
| MSWT Sats Change | 0 (5)<sup>2</sup> | 0 (4.0)<sup>2</sup> | 0.91 |
| Vigilance | 3.8 ± 10.6<sup>1</sup> | 1.8 ± 13.6<sup>1</sup> | 0.85 |
| FEV1/FVC | 0 (0.1)<sup>2</sup> | 0 (0.1)<sup>2</sup> | 0.55 |
| Avoidance – Alone | 0.1 (0.4)<sup>2</sup> | 0.1 (0.2)<sup>2</sup> | 0.91 |
| Avoidance -Accompanied | 0.0 (0.3)<sup>2</sup> | 0.1 (0.2)<sup>2</sup> | 0.55 |
| St George – Active | 6.7 (10.6)<sup>2</sup> | 3.3 (12.8)<sup>2</sup> | 0.55 |
| St George – Impact | 4.3 ± 7.7<sup>1</sup> | 1.5 ± 8.1<sup>1</sup> | 0.55 |
| St George – Symptom | 4.0 ± 11.5<sup>1</sup> | 4.3 ± 10.7<sup>1</sup> | 0.92 |
| Trait | 4.0 (5.5)<sup>2</sup> | 1.0 (7.0)<sup>2</sup> | 0.55 |
| Breathlessness Anxiety (wA) | 0.4 (29.0)<sup>2</sup> | 5.0 (17.8)<sup>2</sup> | 0.15 |
| Breathlessness Severity (wB) | -0.4 ± 21.3<sup>1</sup> | 9.2 ± 23.1<sup>1</sup> | 0.15 |
| Fearful Faces (ms) | 5.2 (116.5) | -13.5 (73.0) |  |
| Happy Faces (ms) | 12.1 (112.8) | -1.9 (106.9) |  |
| Main effect of drug group | F(1,68) = 0.17, p=0.68 |  |  |
| Interaction drug group:emotion | F(1,68) = 0.36, p=0.55 |  |  |

**Table 5.** Scores on questionnaire, behavioural and physiological measures for drug and placebo group following pulmonary rehabilitation (visit three). Measures are expressed as a “change score”. Values are calculated as baseline visit (visit one) minus the later visit, thus positive values indicate an improvement or, in the case of the faces task, a faster reaction time (ms). Significance is reported as Family Wise Error (p<0.05) corrected p-values of the difference in scores between drug and placebo groups at visit three, accounting for scores at visit one. Values are either recorded as mean and standard deviation (1) or as median and interquartile range (2) depending on the normality of distribution. **BMI** = Body Mass Index, **MRC** = Medical Research Council clinical measure of breathlessness, **MSWT =** Modified Shuttle Walk Test, **HR** = Heart rate, **Sats** = Peripheral Oxygen saturation, expressed as a percentage.

|  | Visit Three |  |  |
| --- | --- | --- | --- |
|  | D-cycloserine | Placebo | p-value |
| Catastrophising | 3.0 (7)<sup>2</sup> | 4.5 (8)<sup>2</sup> | 0.86 |
| Depression | 3.1 ± 6.6<sup>1</sup> | 4.7 ± 5.3<sup>1</sup> | 0.86 |
| D12 | 3.0 (8.3)<sup>2</sup> | 2.5 (8)<sup>2</sup> | 0.86 |
| Fatigue | 9.7 ± 12.5<sup>1</sup> | 5.7 ± 12.0<sup>1</sup> | 0.86 |
| BMI | 0.0 (0.8)<sup>2</sup> | -0.1 (0.9)<sup>2</sup> | 0.86 |
| MRC | 0 (1)<sup>2</sup> | 0 (1)<sup>2</sup> | 0.86 |
| MSWT Distance (m) | 40 (92.5)<sup>2</sup> | 30 (70)<sup>2</sup> | 0.86 |
| MSWT Borg Start | 0 (0.5)<sup>2</sup> | 0 (1)<sup>2</sup> | 0.86 |
| MSWT Borg Change | 0.1 ± 1.9<sup>1</sup> | -0.1 ± 1.6<sup>1</sup> | 0.86 |
| MSWT HR Start | 2.7 ± 10.4<sup>1</sup> | 2.4 ± 13.5<sup>1</sup> | 0.98 |
| MSWT HR Change | -51 (29.3)<sup>2</sup> | -53.5 (27)<sup>2</sup> | 0.86 |
| MSWT Sats Start | 0.0 (2)<sup>2</sup> | 0.0 (3)<sup>2</sup> | 0.86 |
| MSWT Sats Change | -1 (7)<sup>2</sup> | 0 (3)<sup>2</sup> | 0.86 |
| Vigilance | 5.0 (12)<sup>2</sup> | 5.5 (12)<sup>2</sup> | 0.86 |
| Spirometry | 0 (0.1)<sup>2</sup> | 0 (0.1)<sup>2</sup> | 0.86 |
| Avoidance – Alone | 0.1 (0.3)<sup>2</sup> | 0.1 (0.3)<sup>2</sup> | 0.98 |
| Avoidance - Accompanied | 0.0 (0.3)<sup>2</sup> | 0.1 (0.3)<sup>2</sup> | 0.86 |
| St George – Active | 6.0 (15.1)<sup>2</sup> | 6.2 (13.4)<sup>2</sup> | 0.86 |
| St George – Impact | 5.1 (12.7)<sup>2</sup> | 5.0 (8.6)<sup>2</sup> | 0.86 |
| St George – Symptom | 6.5 ± 12.5<sup>1</sup> | 6.4 ± 12.7<sup>1</sup> | 0.97 |
| Trait | 2.7 ± 5.8<sup>1</sup> | 3.3 ± 7.6<sup>1</sup> | 0.86 |
| Breathlessness Anxiety (wA) | 3.0 (7.0)<sup>2</sup> | 7.3 (16.8)<sup>2</sup> | 0.053 |
| Breathlessness Severity (wB) | 3.1 (15.6)<sup>2</sup> | 8.4 (13.1)<sup>2</sup> | 0.053 |
| Fearful Faces (ms) | -0.2 (70.7) | -58.2 (88.4) |  |
| Happy Faces (ms) | -10.5 (126.7) | -43.5 (106.4) |  |
| Main effect of drug group | F(1,68) = 0.001, p=0.97 |  |  |
| Interaction drug group:emotion | F(1,68) = 0.002, p=0.97 |  |  |

Overall changes in self-report questionnaires over the course of pulmonary rehabilitation were as expected and are presented in Supplementary Table 5.

## Discussion

### Key Findings

We found that 250mg of D-cycloserine administered prior to the first four sessions of pulmonary rehabilitation showed no effect on breathlessness related brain activity in the 5 regions of interest tested: amygdala, anterior insula, posterior insula, anterior cingulate cortex or hippocampus when assessed either during (after 4-5 sessions) or after the course. Likewise, there was no effect on the secondary endpoints of voxel-wise activity across a wider selection of brain regions. These findings suggest that D-cycloserine does not have the potential to move to phase 3 clinical trials in pulmonary rehabilitation and that alternative drug candidates should be considered.

This is the first study to test a neuro-pharmacological adjunct to pulmonary rehabilitation. A key strength of this study, in addition to its sample size, which is large for a neuroimaging study of breathlessness, is its use of robust clinical trial methodology which was monitored in accordance with GCP standards. We included a formal sample size calculation powered for validated end points relevant to the patient population, pre-registered the study design and published a statistical analysis plan ahead of unblinding. Additionally, our hypotheses focused on five a-priori defined brain regions with demonstrated evidence of strong modulation by D-cycloserine [9, 39] or links to body, symptom perception as well as emotional salience [35, 36]. Together these steps, which are rarely carried out for neuroimaging studies, ensured rigorous methodology, robust findings and provide the best chance to detect an effect if present.

### Why was no drug effect observed?

#### D-cycloserine may not have sufficient glutamatergic activity

While D-cycloserine has not shown sufficient promise to be progressed to full-scale clinical trials, other drugs may now be more attractive candidates. Paired with cognitive therapies as treatment for treatment resistant depression, ketamine and its derivative Esketamine, which blocks pre-synaptic NMDA receptor signalling, increasing glutamate and thereby synaptic plasticity, has been linked with reductions in fear and anxiety, and rapid relief from symptoms [40].

#### Individual differences

Recent literature has highlighted the importance of individual differences in response to D-cycloserine. New evidence suggests that D-cycloserine has the potential to reinforce either positive or negative experiences during exposure based CBT [17]. Direction of action appears to depend on stress levels, which at the neurochemical level influence neurotransmitter concentrations surrounding the NMDA-receptors [41]. These opposing effects may render brain activity differences unobservable in a simple contrast of means. An excellent example can be observed from a reanalysis of two trials which initially found no-overall effect of D-cycloserine. Yet, when post exposure sessional fear was accounted for, the D-cycloserine group evidenced significantly greater clinical improvement than the placebo group. However, this was only the case if fear was reduced. Where fear was high at the end of a session, the D-cycloserine group actually exhibited less clinical improvement than the placebo group [42].

#### Ceiling effect

The action of D-cycloserine is known to be curtailed near the therapeutic ceiling [43], and pulmonary rehabilitation is a highly effective treatment [2], this may leave insufficient scope for improvement in some individuals. There is some evidence to suggest that D-cycloserine speeds up therapeutic effects, even after a single session CBT [6]. To account for the high efficacy of pulmonary rehabilitation, and the potential for D-cycloserine to speed up learning, we also analysed brain activity after the first four rehabilitation sessions. Even at this earlier time point we found no effect of D-cycloserine. This result is in line with Rosenfield et al, who in their meta-analysis, discounted number of treatment sessions from their models after it was found to be non-significant [16].

#### Little evidence of D-cycloserine in older adults

Mechanisms of drug effect may occur via mediated activity within hippocampus, amygdala and insula, regions which overlap with brain networks of internal bodily sensation (interoception) and reappraisal [6, 9, 10]. Given that our previous work, which examined changes in brain activity over a course of pulmonary rehabilitation, identified decreased brain activity within insula correlating with improvements to breathlessness [4], we hypothesised D-cycloserine would work on this body and reappraisal network. However, older adults are not well represented within the evidence base regarding D-cycloserine’s action. Given the well-established changes to NMDA receptor function as the brain ages, D-cycloserine may act differently in this population [44].

#### Alternative brain pathways may be more relevant

Glucocorticoids such as cortisol, combined with exposure-based CBT, have shown promise in reduction of fear in phobias and post-traumatic stress disorder [7] via their action on glucocorticoid receptors along the hypothalamic-pituitary-adrenal (HPA) axis. The exact mechanisms underlying these effects is not yet fully understood, however current evidence points towards either interference with retrieving fearful memories, or enhanced consolidation of the new advantageous memories acquired during extinction [45]. Findings regarding selective serotonin uptake inhibitors (SSRIs) meanwhile are mixed. Paired with CBT, Paroxetine was found to reduce panic attacks by 50% compared to placebo [46]. However, a review of wider SSRI literature found that chronic and sub-chronic administration was associated with reduced CBT response, leading to questions as to whether SSRI’s may even interfere with CBT effectiveness [45]

Collectively these candidate drugs boost synaptic plasticity, although via different neurochemical pathways, which may facilitate the re-setting of fearful associations within the brain. These could be used either during pulmonary rehabilitation, or as part of a precursor programme, helping to recruit harder to reach patients and support self-management. Successful self-management has been highlighted as a key objective by the department of health [47] and often follows on from CBT-based programmes such as the talking therapies recommended by the British Lung Foundation

#### Dosing and dose timing

Questions do still remain regarding D-cycloserine’s optimum dosage, dose timing and number of administered sessions [16]. One recent literature review suggests that up to nine doses, delivered 60-minutes before exposure may yield the best results and increase effect size by up to 50% [16]. Based on the available literature at the time [6, 8] and practical considerations regarding drug availability, we selected a dose of 250mg for this study, administered at the first four rehabilitation sessions. However, given that our maximum effect size was 0.18 (Figure 3), even a 50% increase would be below the commonly reported effect sizes of 0.4-0.7, and up to 1.06 [10, 13, 14, 19]. This strongly suggests that dose and dose timing did not drive the negative result.

**Figure 3.**
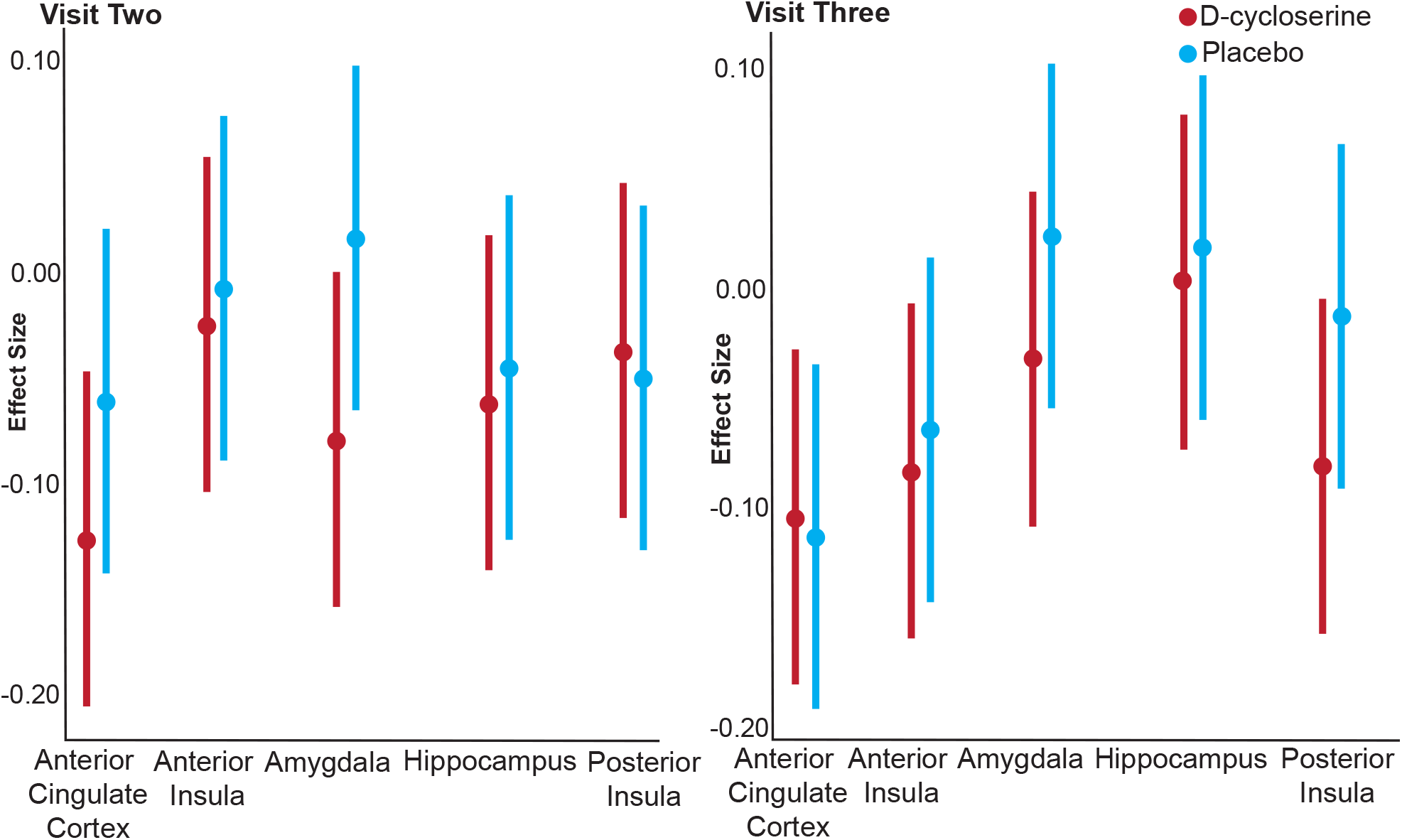
Effect sizes and confidence intervals for each of the five regions of interest at visits two and three for D-cycloserine and Placebo groups.

## Conclusions

We have shown evidence that D-cycloserine does not have an effect on breathlessness related brain activity, behavioural or physiological measures over the course of pulmonary rehabilitation. This study contributes important information regarding the (un)suitability of D-cycloserine as a candidate for phase 3 clinical trials in pulmonary rehabilitation.

## Data Availability

The research materials supporting this publication can be accessed by contacting

## Acknowledgements

The authors would like to thank Jane Francis and the radiography team at the Oxford Centre for Magnetic Resonance Imaging; Beverly Langford, Eleanor Evans and Anja Hayen for their assistance with data collection; Oxford, Reading and Milton Keynes pulmonary rehabilitation teams with particular thanks to Emma Tucker and Chris Swindale, Katy Beckford and Catherine Darby, Milan Bhattacharya and Louise Worrall and the Oxford Respiratory Trials Unit.

## Notes

**Funding** This work was supported by the Dunhill Medical Trust (Grant R333/0214) and the National Institute for Health Research Biomedical Research Centre (Grant RCF18/002) based at Oxford University Hospitals NHS Foundation Trust and The University of Oxford. This research was funded in whole, or in part, by the Wellcome Trust 203139/Z/16/Z. For the purpose of Open Access, the author has applied a CC BY public copyright licence to any Author Accepted Manuscript version arising from this submission. OKH was supported via funding from the European Union’s Horizon 2020 research and innovation programme under the Grant Agreement No 793580, and as a Rutherford Discovery Postdoctoral Fellow from the Royal Society of New Zealand. CJH is supported by the National Institute for Health Research Biomedical Research Centre based at Oxford Health NHS Foundation Trust and The University of Oxford, and by the UK Medical Research Council. KTSP and NMR are supported by the National Institute for Health Research Biomedical Research Centre based at Oxford University Hospitals NHS Foundation Trust and the University of Oxford. AR is funded by a fellowship from MQ. Transforming mental health.

**Disclosures** Dr. Harmer has valueless shares in p1vital and serves on their advisory panel. She has received consultancy payments from p1vital, Zogenix, J&J, Pfizer, Servier, Eli-Lilly, Astra Zeneca, Lundbeck. Dr. Pattinson and Dr Ezra are named as co-inventors on a provisional U.K. patent application titled “Use of cerebral nitric oxide donors in the assessment of the extent of brain dysfunction following injury. Dr. Rahman, has received consulting fees from Rocket Medical U.K. Prof. Nichols has received consulting fees from Perspectum Diagnostics. The remaining authors have no biomedical financial interests or potential conflicts of interest.

### Competing Interest Statement

The authors have declared no competing interest.

### Clinical Trial

NCT01985750

### Funding Statement

This work was supported by the Dunhill Medical Trust (Grant R333/0214) and the National Institute for Health Research Biomedical Research Centre (Grant RCF18/002) based at Oxford University Hospitals NHS Foundation Trust and The University of Oxford. This research was funded in whole or in part by the Wellcome Trust 203139/Z/16/Z. For the purpose of Open Access the author has applied a CC BY public copyright licence to any Author Accepted Manuscript version arising from this submission. OKH was supported via funding from the European Unions Horizon 2020 research and innovation programme under the Grant Agreement No 793580 and as a Rutherford Discovery Postdoctoral Fellow from the Royal Society of New Zealand.
CJH is supported by the National Institute for Health Research Biomedical Research Centre based at Oxford Health NHS Foundation Trust and The University of Oxford and by the UK Medical Research Council. KTSP and NMR are supported by the National Institute for Health Research Biomedical Research Centre based at Oxford University Hospitals NHS Foundation Trust and the University of Oxford. AR is funded by a fellowship from MQ. Transforming mental health.

### Author Declarations

Study approval was granted by South Central Oxford REC B (Ref: 118784, Ethics number: 12/SC/0713).

### Summary of Updates

Manuscript is restructured to contain only primary and secondary analysis

