## Supplementary materials for "The effect of D-cycloserine on brain processing of breathlessness over pulmonary rehabilitation - an experimental medicine study"

Olivia K. Harrison

Sara Booth

Andrea Dennis

Martyn Ezra

Catherine J. Harmer

Mari Herigstad

Bryan Guillaume

Thomas E. Nichols

Najib Rahman

Andrea Reinecke

Olivier Renaud

Kyle T.S. Pattinson

**Online Data Supplement**

***Table 1.*** *Demographic information from the 91 participants who were randomised. Variance is expression either in terms of standard deviation (SD) or interquartile range (IQR).* ***BMI*** *= Body Mass Index,* ***MRC*** *= Medical Research Council clinical measure of breathlessness.* ***$SpO_2$%*** *= Peripheral Oxygen saturation, expressed as a percentage.*

| **Visit 1 (N=91)** | Total | D-cycloserine | Placebo |
|---|---|---|---|
| Age (median years/range) | **70 / (46-85)** | **70 / (47-81)** | **71.5 / (46-85)** |
| Smoking pack-years (IQR) | **30 / (28.5)** | **30 / (25.1)** | **30 /(31.0)** |
| BMI kg.m-2 ± SD | **27.5 ± 5.4** | **26.9 ± 5.5** | **28.1 ± 5.3** |
| MRC (IQR) | **3 (1)** | **3 (1)** | **3 (1)** |
| Resting $SpO_2$% (IQR) | **95 / (3.0)** | **95 / (3.8)** | **95 / (3.0)** |
| Resting heart rate beats.min-1 ± SD | **81.7 ± 13.9** | **82.3 ± 13.4** | **81.0 ± 14.6** |
| FEV1/FVC (IQR) | **0.55 / (0.21)** | **0.54 / (0.24)** | **0.58 / (0.11)** |

#### Randomisation Procedure

Study drugs were purchased from Ipswich Hospital Pharmacy Manufacturing Unit, Heath Road, Ipswich IP4 5PD, Tel: 01473 703603.

Once the participant gave written consent to the trial and completed the MRI scan, a member of the team submitted a randomisation form, entering eligibility criteria and minimisation factors. Allocation to active or placebo capsules, which were both over-encapsulated to appear identical, was carried out by Sealed Envelope Randomisation Services (Sealed Envelope Ltd, Concorde House, Grenville Place, London NW7 3SA). The randomisation number was then provided to the Oxford Respiratory Trials Unit who dispensed the drug/placebo. Randomisation codes were held by Sealed Envelope until study completion, after which at the first stage of unblinding an independent researcher provided study researchers with a coded binarised system for analysis. Researchers remained blinded to group identity until analysis was completed.

**Sample Size**

At the time of study inception (and to a large extent still to date), the literature regarding D-cycloserine's effects on functional brain activity is very limited. Therefore, in order to calculate the sample sizes required for this study we first took into account the described effects of D-cycloserine in clinical studies of augmentation for cognitive behavioural therapy for anxiety disorders, where effect sizes of up to 1.06 have been reported (although more commonly 0.4 to 0.7) [1-4]**.** The most relevant paper (on treatment of snake phobia [5]**)** demonstrated that effects observed with neuroimaging were more sensitive than behavioural effects, therefore we believe that powering for a behavioural outcome measure (breathlessness-anxiety) provided a safe margin and was likely to be sufficiently conservative to detect our measures of interest. This was particularly the case as compared to the relatively blunt nature of behavioural data collection, functional neuroimaging carries considerably more specificity and statistical power. The study was not therefore specifically powered to investigate the clinical effects of D-cycloserine. In our previous study we observed an 11% (SD15%) improvement in breathlessness-related anxiety, measured with our FMRI word task (pre-treatment mean score 38%, post treatment mean score 27%, difference 11%, SD of difference 15%) [6]. Making a conservative assumption, we estimated that D-cycloserine augments this response with an effect size of 0.4. Assuming a similar coefficient of variation we anticipated an 18% (SD24%) improvement in breathlessness-anxiety (i.e. pre-treatment mean score 38%, post treatment mean score 20%, difference 18%, SD of difference 24%). Assuming $\alpha=0.05$ and power 0.80, then we estimated a sample size of 36 in each group randomised 1:1. As this is a behavioural outcome, we expected this to have sufficient power to detect change in BOLD signalling.

**Missing Data**

The potential effect of missing brain imaging data was explored using a sensitivity analysis. Missing questionnaire and physiology data points were imputed using the Markov chain Monte Carlo method (multiple imputation technique) within the MICE package in R. A summary of missing data is provided below.

**Behavioural Measures**

*Questionnaire Measures*

**Dyspnoea-12 (D12) Questionnaire**: This is a 12-item questionnaire designed to measure the severity of breathlessness and has been validated for use in patients with respiratory disease [7].

**Centre for Epidemiologic Studies Depression Scale (CES-D)**: Depressive symptoms are commonly observed in patients with respiratory disease. This brief questionnaire consists of 20 items investigates the symptoms of depression across a number of factors [8].

**State-Trait Anxiety Inventory (STAIT-T)**: This questionnaire assesses participant's general level of anxiety in particular scenarios via 20 questions asking "how anxious you generally feel" [9].

**Fatigue Severity Scale**: This 9-point questionnaire quantifies patient fatigue, which is well documented in its association with COPD [10].

**St George's Respiratory Questionnaire (SGRQ)**: There are 50 questions in this questionnaire, which has been developed and validated for use in COPD and asthma. The questions measure the impact of overall health, daily life and well-being [11].

**Medical Research Council (MRC) breathlessness scale**: The MRC scale quantifies perceived difficulty due to respiratory restrictions on a scale of 1 to 5 [12].

**Mobility Inventory (MI)**: This questionnaire collects data regarding the extent to which a participant avoids certain situations, either alone or accompanied (21-items in each category) [13].

**Breathlessness Catastrophising Scale –** adapted from the catastrophic thinking scale in asthma: This 13-point questionnaire was modified for this study by substituting the word "asthma" for "breathlessness" in order to measure catastrophic thinking [14] [15].

**Breathlessness Vigilance Scale –** adapted from the pain awareness and vigilance scale: This questionnaire was modified by substituting the word "breathlessness" for the word "pain". The 16-point scale measures how much a participant focuses their attention onto their breathlessness [16] [15].

**Physiological Measures**

A trained respiratory nurse collected spirometry measures of $FEV_1$ and FVC using Association for Respiratory Technology and Physiology standards [17]. Participants performed two modified incremental shuttle walk tests (MSWT) [18], and heart rate and oxygen saturations ($SpO_2$) were measured immediately before the MSWT and subsequently every minute until 10 minutes post-exercise (or until participants returned to their baseline state) using a fingertip pulse oximeter ($Go_2$; Nonin Medical Inc). Before and after the MWST participants also rated their breathlessness on a modified Borg scale [19]. In a MWST participants must walk between and around two cones, placed 10m apart in time to a set of auditory beeps played from a laptop. Initially the speed of beep repetition is slow, but the participant must increase their walking speed each minute in order to reach the cone before the next beep. Participants continue to walk (or run) until they are too breathless to continue, at which point the total distance walked is recorded.

**MRI Acquisition**

Prior to each MRI session participants were screened for standard MRI contraindications including metal in or about their person, epilepsy and claustrophobia.

*Image acquisition:*

Hardware: A Tim System (Siemens Healthcare GmbH) 12-channel head coil.

T1 sequence parameters: TR, 2040ms; TE, 4.68ms; voxel size, 1 x 1 x 1 mm; FOV, 200mm; flip angle, 8°; inversion time, 900ms; bandwidth 130 Hz/Px).

T2*-weighted (functional) sequence parameters: TR, 3000ms; TE 30ms; voxel size 3 x 3 x 3 mm; FOV, 192mm; flip angle 87°; echo spacing 0.49ms.

Functional scan durations: word-task - 215 volumes, 7 minutes and 33 seconds duration and faces task - 168 volumes, 5 minutes and 42 seconds duration.

Field map scans of the $B_0$ field were obtained to aid the distortion correction of the functional scans: TR, 488ms; TE1, 5.19ms; TE2, 7.65ms; flip angle 60°; voxel size, 3.5 x 3.5 x 3.5 mm.

*Word Task*

This task was developed and published by Herigstad and colleagues in 2016 for use in the COPD population [20]. Word cues were developed in three key stages; firstly in collaboration with respiratory practitioners, academics and physiotherapists, a set of 30 word cues associated with breathlessness were created. Next, these cues were provided to patients with COPD alongside a VAS rating scale, allowing patients to rate how breathless and anxious the situations identified by the cues would make them feel. Following adjustments based on participant feedback, the word cues were then computerised and tested in a larger population of COPD patients [20]. Further validation was carried out in the fMRI environment and by for clinical sensitivity with comparisons between changes in key questionnaire measures and word-cue rating.

Before the first scan session, participants were given the opportunity to practice using the button box with a set of test words.

Control tasks

1) A control condition, used as a baseline measure of activity in response to the presentation of a visual stimulus was presented 4 times over the course of the word-cue scan, consisting of a string of “XXXXXXXXXXXXXXX” with fixed length of 15 characters, and each time was presented for 7 seconds. No rating period followed these control blocks [6, 15, 20].

2) A validated task of emotional faces was used as a control to separate generalized anxiety from breathlessness specific anxiety. Emotional facial expressions are widely recognised to activate the same brain pathways as the behavioural emotion conveyed by the expression itself. Fearful facial expressions, for example, have been shown to correspond to activity within the amygdala, a region known to modulate fear processing [26]. Faces were drawn from a set first developed by Ekman and Friesen [21] and furthered by Young et al [22]. Photographs of 10 faces (5 male, 5 female) with fearful or happy expressions of 100% intensity were used. Each face was shown for 500ms in blocks of 30 seconds. A fixation cross was interspersed for 30 seconds between the blocks of faces. Participants were instructed to respond via a button box to indicate facial gender. Reaction time and accuracy were recorded throughout the task. The task contrasting fear and happy facial expressions has been extensively used in previously studies and has been found to activate the amygdala in both healthy volunteers and in depressed patients [23, 24]. Neutral faces are not typically used as they can be interpreted as threatening or ambiguous in different settings [25].

***Imaging Analysis***

*Functional MRI Preprocessing*

Data denoising was carried out as follows: Before the first level analysis, each functional scan was decomposed into maximally independent components using FMRIB's MELODIC tool (Multivariate Exploratory Linear Optimised Decomposition into Independent Components). "Noise" components were identified by FIX (FMRIB's auto-classification tool, [26, 27]) using the WhII.Standard.RData [28] trained classifier with aggressive clean up option. A Principal Component Analysis (PCA) was run on the FIX identified components to retrain 99% of the variance. Separately, the cardiac and respiratory related physiological signals (recorded via a pulse oximeter and respiratory bellows) were transformed into a series of regressors, (three cardiac and four respiratory harmonics) as well as an interaction term and a measure of respiratory volume per unit of time (RVT), using FSL's physiological noise modelling tool (PNM). The signal associated with these waveforms (modelled using retrospective image correction (RETROICOR) [29, 30]) was then used to form voxelwise noise regressors.

The confounds identified by FSL's FIX and PNM tools, along with sources of noise arising from motion, were then combined into a single model. This single noise model approach builds upon the technique outlined by [31]; and fully detailed by [32]. In these preceding works we employed a step-wise technique whereby physiological noise (identified by PNM) and FIX-identified noise were each removed from the data in separate steps prior to data entry into the lower level model. In the new cleanup pipeline, a single text file containing time-course information relating to FIX identified noise components along with white matter or CSF related noise was included as additional confound EV's within the lower level model, while the PNM-identified noise was entered into the model as a standard voxel-wise confound list. In this updated de-noising pipeline, confounds identified above are added to model at the stage of first-level analysis and thus the functional dataset can be corrected for sources of noise arising from motion, scanner and cerebro-spinal fluid artefacts, cardiac, and respiratory noise in a single step, rather than three.

*Functional MRI Analysis*

MRI processing was performed using FEAT (FMRI Expert Analysis Tool within the FSL package). The data were corrected for movement using MCFLIRT (Motion correction using FMRIB's Linear Image Registration Tool [33]). Non-brain structures were removed using BET (Brain Extraction Tool [34]). Spatial smoothing was carried out using a full-width-half-maximum Gaussian kernel of 5mm, while high-pass temporal filtering (Gaussian-weighted least squares straight line fitting; 90 s) removed low frequency noise and slow-drift. Distortion correct of EPI data was carried out using a combination of FUGUE (FMRIB's Utility for Geometrically Unwarping EPI's [35, 36] and BBR (Boundary Based Registration; part of the FMR Expert Analysis Tool, FEAT version 6.0 [37]). The data were corrected for physiological noise using FSL's FIX-PNM pipeline. Functional scans were registered in a two-step process to the MNI152 (1x1x1 mm) standard space brain template. Firstly, each subject's EPI was registered to their associated T1-weighted structural image using BBR (6 DOF) with nonlinear field map distortion correction [37]. In the second step the subject's structural image was registered to 1mm standard space via an affine transformation followed by nonlinear registration (using FNIRT: FMRIB's Non-linear Registration Tool [38]).

*Region of interest extraction*

The five bilateral regions of interest (ROI) were anterior insula cortex, posterior insula cortex, anterior cingulate cortex, amygdala and hippocampus. Seed voxels for each region of interest were identified as the peak voxel co-ordinates (Supplementary table 2) responding to breathlessness word-cues published by Herigstad et al 2017 [6] within the boundaries of each region of interest identified by standard atlas maps. The seed voxels were expanded to include the surrounding voxels within a 5 mm radius.

Left and right masks of bilateral regions of interests (anterior insula, posterior insula, anterior cingulate, amygdala and hippocampus) were added together to form one mask for each region of interest. Following registration each mask was re-thresholded at 40% probability to avoid interpolation errors before being binarised.

***Table 2.*** *MNI coordinates for region of interest seeds*

| Region of interest | Hemisphere | x | y | z |
|---|---|---|---|---|
| Anterior insula cortex | Left | -31 | 7 | -14 |
| | Right | 38 | 13 | -10 |
| Posterior insula cortex | Left | -31 | 7 | -15 |
| | Right | 37 | 10 | -12 |
| Anterior cingulate cortex | | -5 | 34 | -3 |
| Amygdala | Left | -19 | -7 | -21 |
| | Right | 19 | -8 | -18 |
| Hippocampus | Left | -22 | -12 | -26 |
| | Right | 20 | -9 | -19 |

*Network mask*

A second network mask region of interest was created from the 5 core regions of interest outlined above and an additional 11 regions defined by standard anatomical atlas maps (Harvard-Oxford Atlas and Destrieux' cortical atlas) (Supplementary Figure 1). A 40% probability threshold was applied to each region, before they were combined along with the original 5 regions into one network mask. This network mask was then registered to each individual before being re-thresholded at 40% probability to avoid interpolation errors and binarized. Combining the 16 regions into a single mask enabled us to appropriately correct for multiple comparisons.

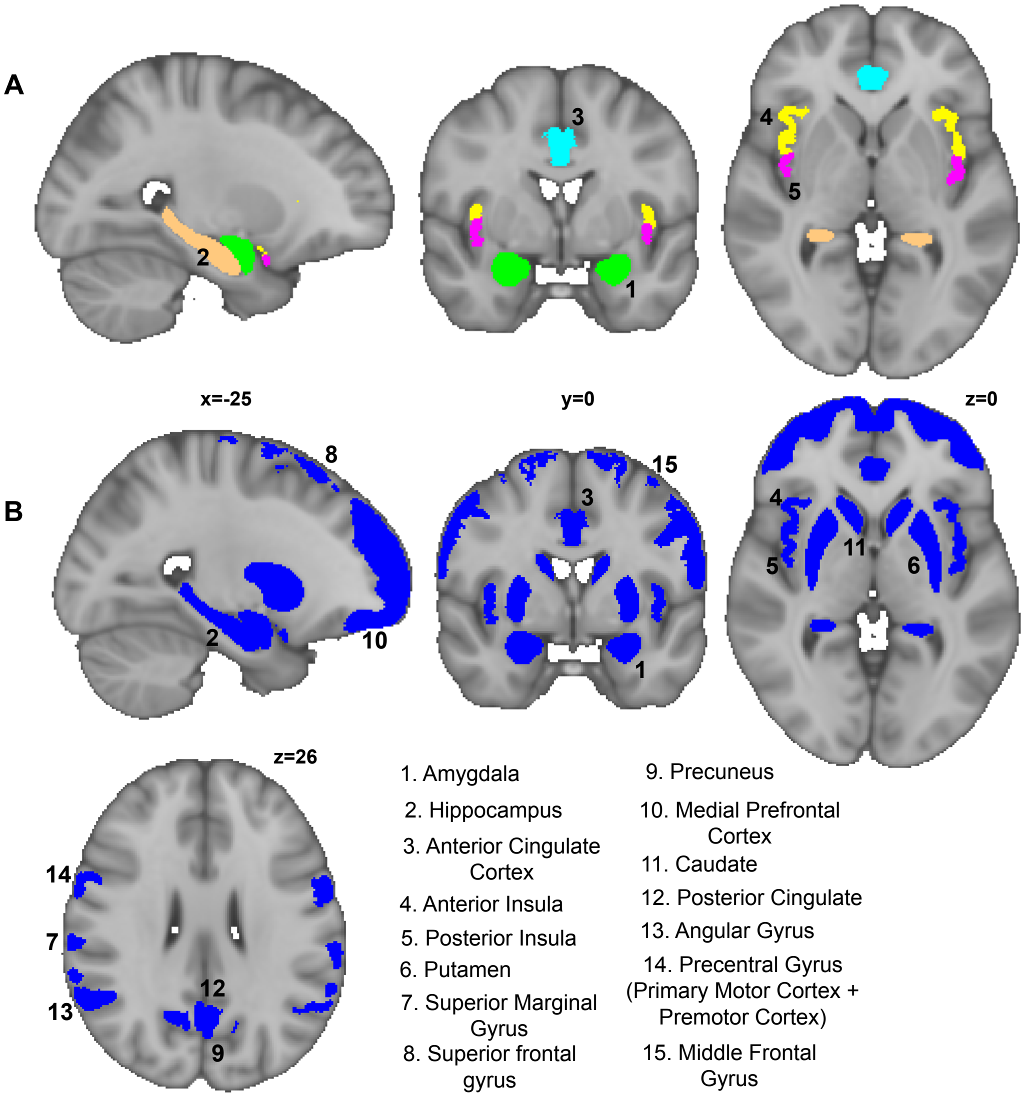


***Figure 1.*** *Panel A highlights the 5 key regions of interest while Panel B shows the expanded region of interest map.*

*Word cue task*

At the individual subject level, a general linear model (GLM) was created with explanatory variables (EVs) for the 7 seconds of word or non-word presentation, and two (7 second) de-meaned EVs modeling the reported breathlessness and anxiety response to the word cues. An additional explanatory noise variable was included to model the period during which the participant responded using the visual analog scale (VAS).

*Control task*

At the individual subject level, a GLM was created with explanatory variables for the 30 second stimulus presentation periods of happy and fearful faces, along with the associated (de-meaned) reaction times. Two additional explanatory variables were created to model participant (de-meaned) accuracy in identifying whether the presented faces were male or female.

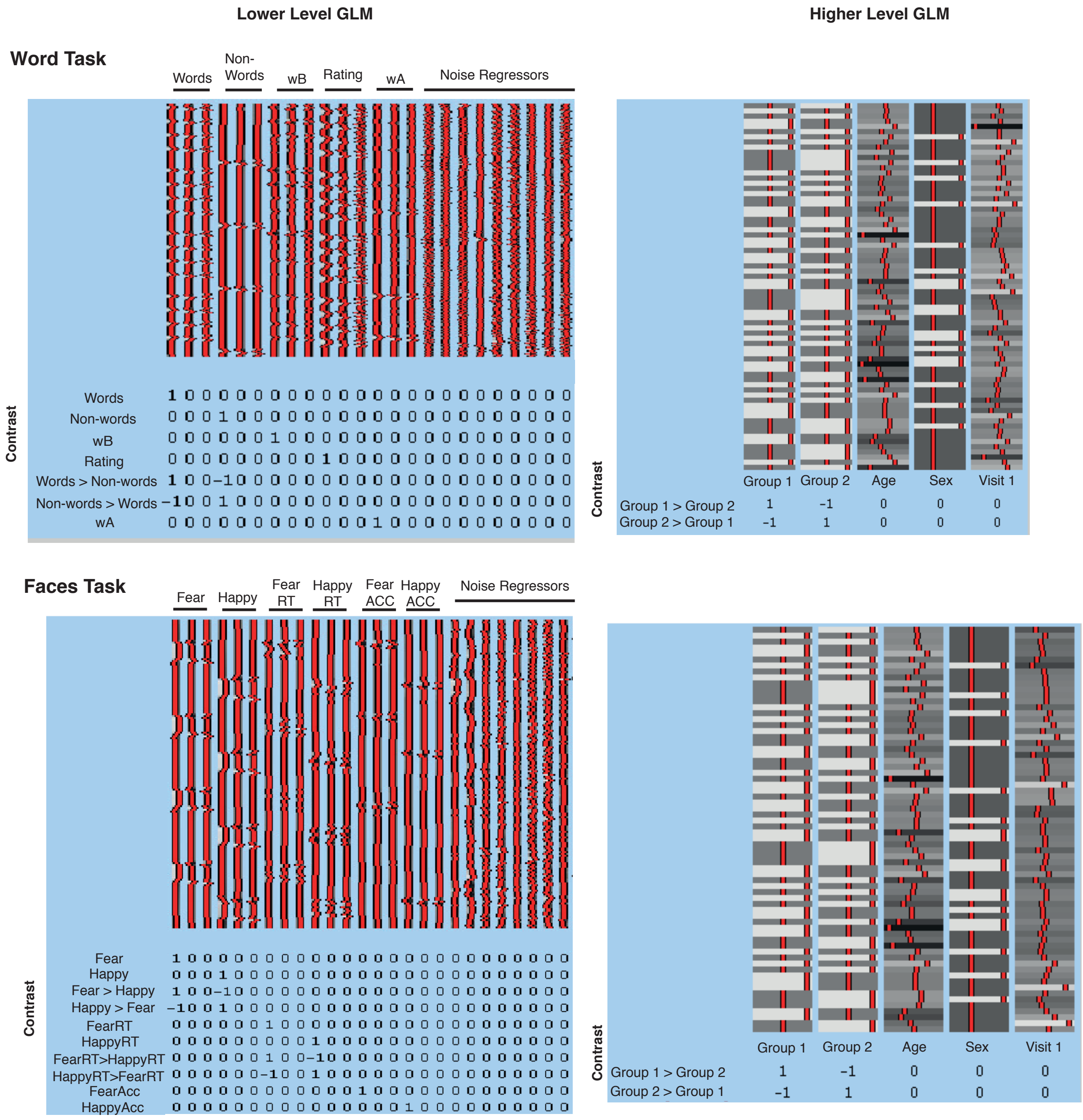


***Supplementary Figure 2** – An illustration of the generalised linear models (GLM) used for both lower and higher level analyses for the word and faces task. Abbreviations as follows – wB – breathlessness rating, wA – breathlessness anxiety rating, RT – reaction time, ACC – accuracy.*

***Statistical Analysis***

*Sensitivity Analysis*

$$\triangle = \triangle_{CC} + Y_1 P_1 - Y_2 P_2$$

**Equation 1.** Formula for calculating the outcome of a range of missing not at random scenarios. Where delta is the treatment effect under the missing-not-at-random. $Delta_{CC}$ is the treatment effect under a complete case scenario. $Y_1$ and $Y_2$ are the assumed mean responses for patients with missing data in treatment groups 1 and 2 respectively. $P_1$ and $P_2$ are the proportion of patients missing in groups 1 and 2 respectively.

Sensitivity analysis essentially asks the question "if all of the participants who were enrolled but did not complete the study actually had, would the result of the primary analysis have changed?" To answer this, a number of different brain activity levels are simulated to account for reasonable extreme scenarios.

Using Equation 1, three quantiles of brain activity within each of the five key regions of interest in response to breathlessness-related word cues were calculated for participants in treatment group 1. This corresponded to 25%, 50% and 75% of activity observed within the group of participants who completed all three visits.

The standard error of delta is approximately equal to the standard error for $delta_{CC}$. $Y_1$ will be varied between $Y_2$ – (5%) and $Y_2$ + (5%). These values were multiplied by the proportion of missing participants from each group. The simulated complete datasets were entered into linear mixed effects models where they were adjusted for age and gender. To correct to multiple comparisons across regions, permutation testing (with Family Wise Error Rate (FWE) 5%) was carried out.

### Results

#### Behavioural

#### Missing data

***Table 3.*** *Missing data reported as the total percentage of data collected for that measure.* ***MSWT –*** *modified shuttle walk test;* ***HR –*** *Heart rate;* ***Sats –*** *Oxygen saturation;* ***BMI –*** *Body mass index;* ***MRC –*** *Medical research council breathlessness scale*

| | **Pre-rehabilitation** | | **During rehabilitation** | | **Following rehabilitation** | |
|---|---|---|---|---|---|---|
| | D-cycloserine | Placebo | D-cycloserine | Placebo | D-cycloserine | Placebo |
| **MSWT** | | | | | | |
| Distance | **11** | **9** | **14** | **3** | **14** | **9** |
| Borg change | **11** | **9** | **14** | **3** | **14** | **9** |
| HR change | **11** | **9** | **14** | **3** | **14** | **9** |
| Sats change | **11** | **9** | **14** | **3** | **14** | **9** |
| BMI | **-** | **-** | **8** | **3** | **30** | **21** |
| MRC | **-** | **-** | **-** | **-** | **3** | **3** |

***Table 4.*** *Scores on questionnaire, physiology and behavioural measures for drug and placebo group before (visit one), during (visit two) and after (visit three) pulmonary rehabilitation. Variance is expressed either in terms of [1]standard deviation (SD) or [2]interquartile range (IQR).* ***BMI*** *= Body Mass Index,* ***MRC*** *= Medical Research Council clinical measure of breathlessness,* ***MSWT =*** *Modified Shuttle Walk Test,* ***HR*** *= Heart rate,* ***Sats*** *= Peripheral Oxygen saturation, expressed as a percentage*

| | Visit One | | Visit Two | | Visit Three | |
|---|---|---|---|---|---|---|
| | **D-cycloserine** | **Placebo** | **D-cycloserine** | **Placebo** | **D-cycloserine** | **Placebo** |
| Catastrophising | **6.0 (9.3) [2]** | **8.0 (13.0) [2]** | **5.0 (7.2) [2]** | **5.5 (14.0) [2]** | **3.0 (5.0) [2]** | **5.0 (13.0) [2]** |
| Depression | **10.0 (9.0) [2]** | **13.0 (8.0) [2]** | **9.0 (9.3) [2]** | **9.5 (12.0) [2]** | **6.0 (10.3) [2]** | **7.0 (12.0) [2]** |
| D12 | **11.0 (11.3) [2]** | **9.5 (10.0) [2]** | **6.0 (6.5) [2]** | **5.5 (10.0) [2]** | **5.0 (7.0) [2]** | **5.5 (10.0) [2]** |
| Fatigue | **43.0 (19.3) [2]** | **33.5 (25.0) [2]** | **35.0 (23.5) [2]** | **32.5 (27.0) [2]** | **29(18.0) [2]** | **27.5 (18.0) [2]** |
| BMI | **27.3 (6.5) [2]** | **26.9 (5.7) [2]** | **27.3 ± 4.9[1]** | **27.4 ± 4.4[1]** | **27.7 ± 4.8[2]** | **27.0 ± 4.5[1]** |
| MRC | **3 (1.0) [2]** | **3 (1.0) [2]** | **3 (1.0) [2]** | **2 (1.0) [2]** | **2 (1.3) [2]** | **2 (1.0) [2]** |
| MSWT Distance (m) | **280 (291) [2]** | **325 (230) [2]** | **290 (268)** | **355 (250) [2]** | **340 (293) [2]** | **340 (250) [2]** |
| MSWT Borg Start | **0.5 (1.0) [2]** | **0.5 (1.0) [2]** | **0.5 (0.5) [2]** | **0.5 (1.0) [2]** | **0.5 (0.6) [2]** | **0.5 (1.0) [2]** |
| MSWT Borg Change | **3.0 (1.5) [2]** | **2.8 (2.5) [2]** | **2.5 (1.5) [2]** | **3.0 (1.0) [2]** | **3.0 (2.0) [2]** | **2.8 (1.5) [2]** |
| MSWT HR Start | **80.8 ± 13.4[1]** | **80.8 ± 14.9[1]** | **81.1 ± 14.8[1]** | **79.4 ± 12.9[1]** | **78.2 ± 13.1[1]** | **78.4 ± 13.1[1]** |
| MSWT HR Change | **24.0 (27.3)[2]** | **31.5 (16.0)[2]** | **31.0 (19.5) [2]** | **32.5 (28) [2]** | **31.0 (23.5) [2]** | **31.5 (25.0) [2]** |
| MSWT Sats Start | **95.0 (3.3) [2]** | **94.5 (3.0) [2]** | **95.0 (4.0) [2]** | **95.0 (3.0) [2]** | **95.0 (2.5) [2]** | **94.0 (5.0) [2]** |
| MSWT Sats Change | **4 (6.3) [2]** | **5 (5.0) [2]** | **4 (4.3) [2]** | **5 (7.0) [2]** | **6 (8.3) [2]** | **5 (5.0) [2]** |
| Vigilance | **37.0 ± 15.0[1]** | **33.7 ± 17.7[1]** | **33.2 ± 11.1[1]** | **31.9 ± 13.8[1]** | **33 ± 20.3[1]** | **26.5 ± 27[1]** |
| FEV1/FVC | **0.53 ± 0.17[1]** | **0.56 ± 0.13[1]** | **0.43 (0.36) [2]** | **0.60 (0.12) [2]** | **0.50 (0.34) [2]** | **0.60 (0.22) [2]** |
| Avoidance – Alone | **1.5 (0.63) [2]** | **1.5 (0.8) [2]** | **1.4 (0.43) [2]** | **1.4 (0.6) [2]** | **1.4 (0.46) [2]** | **1.4 (0.65) [2]** |
| Avoidance -Accompanied | **1.30 (0.53) [2]** | **1.40 (0.6) [2]** | **1.24 (0.36) [2]** | **1.20 (0.33) [2]** | **1.24 (0.44) [2]** | **1.24 (0.43) [2]** |
| St George – Active | **67.6 ± 19.0[1]** | **57.1 ± 22.8[1]** | **62.6 18.4[1]** | **55.2 19.5[1]** | **61.1 (22.3) [2]** | **52.6 (19.5) [2]** |
| St George – Impact | **32.7 ± 15.8[1]** | **29.3 ± 16.4[1]** | **24.8 (22.7) [2]** | **23.2 (25.7) [2]** | **22.4 (24.8) [2]** | **20.9 (25.4) [2]** |
| St George – Symptom | **60.7 ± 18.5[1]** | **62.9 ± 18.5[1]** | **56.7 ± 1.4[1]** | **58.6 ± 21.2[1]** | **54.2 ± 21.5[1]** | **56.4 ± 20.2[1]** |
| Trait | **36.7 ± 9.9[1]** | **38.5 ± 9.3[1]** | **33.0 (12.3) [2]** | **37.5 (16.0) [2]** | **31.0 (14.8) [2]** | **34.0 (16.0) [2]** |
| Breathlessness Anxiety (wA) | **10.2 (28.9)[1]** | **27.8 (38.6)[1]** | **9.5 (34.7)[1]** | **9.5 (31.4)[1]** | **3.6 (32.7)[1]** | **6.4 (36.8)[1]** |
| Breathlessness Severity (wB) | **43.9 (16.1)[1]** | **51.5 (24.1)[1]** | **44.5 (19.5)[1]** | **42.2 (31.3)[1]** | **40.0 (21.1)[1]** | **41.8 (27.1)[1]** |
| Fearful Faces (ms) | **739.5 (179.5)[1]** | **757.6 (218.8)[1]** | **773.9 (196.0)[1]** | **796.1 (189.2)[1]** | **767 (165.5)[1]** | **837.7 (192.0)[1]** |
| Happy Faces (ms) | **794.7 (174.6)[1]** | **774.8 (201.4)[1]** | **753.8 (151.8)[1]** | **778.6 (161.0)[1]** | **759.3 (178.0)[1]** | **808.3 (142.0)[1]** |

***Table 5.*** *Mean change scores on questionnaire and behavioural measures across both drug and placebo groups following four sessions of pulmonary rehabilitation (visit two). Measures are expressed as a "change score", where visit two scores were subtracted from visit one. Variance is expressed either in terms of [1]standard deviation (SD) or [2]interquartile range (IQR). Significance is reported as exploratory uncorrected p-values and as Family Wise Error (p<0.05) corrected p-values.*

| | Visit Two | | |
|---|---|---|---|
| | **Cohort Change** | **Uncorrected p-values** | **Corrected p-values** |
| **Catastrophising** | 2.0 (5) [2] | p=0.08 | p=0.097 |
| **Depression** | 2.2 ± 6.0 [1] | **p=0.004*** | p=0.06 |
| **D12** | 3.0 ± 5.1 [1] | **p<0.001*** | **p<0.001*** |
| **Fatigue** | 6.0 (11) [2] | **p=0.04*** | p=0.65 |
| **BMI** | -0.04 (0.6) [2] | p=0.71 | p=0.97 |
| **MRC** | 0.0 (0) [2] | p=0.71 | p=0.97* |
| **MSWT Distance** | 0.0 (95.0) [2] | p=0.70 | p=0.97 |
| **MSWT Borg Start** | 0.0 (1) [2] | p=0.63 | p=0.97 |
| **MSWT Borg Change** | 0.5 (2) [2] | p=0.74 | p=0.97 |
| **MSWT HR Start** | 0.5 ± 13.5 [1] | p=0.75 | p=0.97 |
| **MSWT HR Change** | -46 ± 24 [1] | **p<0.001*** | **p<0.001*** |
| **MSWT Sats Start** | -0.2 ± 2.1 [1] | p=0.42 | p=0.97 |
| **MSWT Sats Change** | 0.0 (4) [2] | p=0.97 | p=0.97 |
| **Vigilance** | 2.8 ± 12.1 [1] | p=0.05 | p=0.78 |
| **FEV1/FVC** | -0.01 (0.1) [2] | p=0.75 | p=0.97 |
| **Avoidance -** Alone | 0.1 (0.3) [2] | p=0.19 | p=0.97 |
| **Avoidance -** Accompanied | 0.05 (0.3) [2] | p=0.47 | p=0.97 |
| **St George -** Active | 5.8 (12.0) [2] | p=0.27 | p=0.97 |
| **St George –** Impact | 2.9 ± 7.9 [1] | **p=0.003*** | p=0.05 |
| **St George –** Symptom | 4.2 ± 11.1 [1] | **p=0.002*** | **p=0.04*** |
| **Trait** | 2.0 (6) [2] | p=0.39 | p=0.97 |

***Table 6.*** *Mean change scores on questionnaire and behavioural measures across both drug and placebo groups following pulmonary rehabilitation (visit three). Measures are expressed as a "change score", where visit three scores were subtracted from visit one. Variance is expressed either in terms of [1]standard deviation (SD) or [2]interquartile range (IQR). Significance is reported as exploratory uncorrected p-values and as Family Wise Error (p<0.05) corrected p-values.*

| | Visit Three | | |
|---|---|---|---|
| | **Cohort Change** | **Uncorrected p-values** | **Corrected p-values** |
| **Catastrophising** | 3.0 (8) [2] | **p=0.001*** | **p=0.02*** |
| **Depression** | 3.9 ± 6.0 [1] | **p<0.001*** | **p<0.001*** |
| **D12** | 3.0 (7.6) [2] | **p<0.001*** | **p=0.004*** |
| **Fatigue** | 7.8 ± 12.4 [1] | **p<0.001*** | **p<0.001*** |
| **BMI** | -0.04 (0.9) [2] | p=0.66 | p=0.99 |
| **MRC** | 0.0 (1) [2] | **p=0.01*** | p=0.19 |
| **MSWT Distance** | 30 (80.0) [2] | p=0.32 | p=0.99 |
| **MSWT Borg Start** | 0.0 (0.88) [2] | p=0.38 | p=0.99 |
| **MSWT Borg Change** | -0.01 ± 1.75 [1] | p=0.95 | p=0.99 |
| **MSWT HR Start** | 2.6 ± 11.9 [1] | p=0.07 | p=0.84 |
| **MSWT HR Change** | -51 (27.8) [2] | **p<0.001*** | **p<0.001*** |
| **MSWT Sats Start** | 0.0 (3) [2] | p=0.65 | p=0.99 |
| **MSWT Sats Change** | -1.0 (5.8) [2] | p=0.27 | p=0.99 |
| **Vigilance** | 5.0 (11.8) [2] | p=0.11 | p=0.97 |
| **FEV1/FVC** | -0.01 (0.1) [2] | p=0.99 | p=0.99 |
| **Avoidance - Alone** | -0.1 (0.3) [2] | p=0.10 | p=0.97 |
| **Avoidance - Accompanied** | 0.05 (0.3) [2] | p=0.12 | p=0.97 |
| **St George - Active** | 6.1 (14.0) [2] | p=0.08 | p=0.84 |
| **St George – Impact** | 5.1 (10.9) [2] | **p=0.04*** | p=0.46 |
| **St George – Symptom** | 6.5 ± 12.5 [1] | **p<0.001*** | **p<0.001*** |
| **Trait** | 3.0 ± 6.6 [1] | **p<0.001*** | **p<0.005*** |

### Control Task

***Table 7.*** *Median change in reaction times (ms) in response to fearful or happy faces for both drug and placebo group following pulmonary rehabilitation (visit 3). Measures are expressed as a "change score", where visit 3 scores were subtracted from visit 1. Variance is expressed as interquartile range (IQR). Significance set at exploratory uncorrected p-values and as Family Wise Error (p<0.05) corrected p-values.*

| | Cohort Average | Uncorrected p-values | Corrected p-values |
|---|---|---|---|
| Fearful | **-19.8 (104.2)** | **p=0.28** | **p=0.37** |
| Happy | **-30.4 (102.8)** | **p=0.37** | **p=0.37** |

### Word Task

***Table 8.*** *Rating scores for breathlessness related anxiety (wA) and breathlessness (wB) for both drug and placebo group following pulmonary rehabilitation (visit 3). Measures are expressed as a "change score", where visit 3 scores were subtracted from visit 1. Variance is expressed in terms of interquartile range (IQR). Significance set at exploratory uncorrected p-values and as Family Wise Error (p<0.05) corrected p-values.*

| | Cohort Average | Uncorrected p-values | Corrected p-values |
|---|---|---|---|
| wA | **5.3 (10.0)** [2] | **p=0.01** | **p=0.02** |
| wB | **6.3 (14.9)** [2] | **p=0.07** | **p=0.07** |

### Sensitivity Analysis and Study completeness

Ten participants who were randomised to the D-cycloserine group and eleven participants who were randomised to the placebo group did not complete all three visits. Sensitivity analysis revealed that the inclusion of the 21 missing participants would not have altered the primary outcome.

***Table 9.*** Results of sensitivity analysis for visit two. The significance of linear mixed effects models applied to the simulated complete datasets are reported as Family Wise Error Rate (FWE) 5% corrected p-values for each stimulate quantile.

| Quantile | Region of interest | | | | |
|---|---|---|---|---|---|
| | Anterior cingulate cortex | Anterior insula | Amygdala | Hippocampus | Posterior insula |
| Lower 25% | 0.58 | 0.96 | 0.12 | 0.83 | 0.96 |
| Lower 50% | 0.42 | 0.99 | 0.13 | 0.99 | 0.99 |
| Lower 75% | 0.35 | 0.82 | 0.31 | 0.82 | 0.76 |
| Upper 25% | 0.81 | 0.97 | 0.31 | 0.99 | 0.99 |
| Upper 50% | 0.56 | 0.98 | 0.26 | 0.91 | 0.91 |
| Upper 75% | 0.61 | 0.80 | 0.65 | 0.65 | 0.65 |

***Table 10.*** Results of sensitivity analysis for visit three. The significance of linear mixed effects models applied to the simulated complete datasets are reported as Family Wise Error Rate (FWE) 5% corrected p-values for each stimulate quantile.

| Quantile | Region of interest | | | | |
|---|---|---|---|---|---|
| | Anterior cingulate cortex | Anterior insula | Amygdala | Hippocampus | Posterior insula |
| Lower 25% | 0.72 | 0.72 | 0.06 | 0.27 | 0.10 |
| Lower 50% | 0.53 | 0.39 | 0.08 | 0.40 | 0.19 |
| Lower 75% | 0.73 | 0.26 | 0.17 | 0.73 | 0.46 |
| Upper 25% | 0.98 | 0.96 | 0.29 | 0.70 | 0.29 |
| Upper 50% | 0.83 | 0.65 | 0.28 | 0.83 | 0.48 |
| Upper 75% | 0.90 | 0.43 | 0.40 | 0.91 | 0.66 |
